# Hydrating the Respiratory Tract: An Alternative Explanation Why Masks Lower Severity of COVID-19 Disease

**DOI:** 10.1101/2020.12.23.20248671

**Authors:** Joseph M. Courtney, Ad Bax

## Abstract

Seasonality of respiratory diseases has been linked, among other factors, to low outdoor absolute humidity and low relative humidity in indoor environments, which increase evaporation of water in the mucosal layer lining the respiratory tract. We demonstrate that normal breathing results in an absorption-desorption cycle inside facemasks, where super-saturated air is absorbed by the mask fibers during expiration, followed by evaporation during inspiration of dry environmental air. For double-layered cotton masks, which have considerable heat capacity, the temperature of inspired air rises above room temperature, and the effective increase in relative humidity can exceed 100%. We propose that the recently reported, disease-attenuating effect of generic facemasks is dominated by the strong humidity increase of inspired air.

**SIGNIFICANCE STATEMENT:** Facemasks are the most widely used tool for mitigating the spread of the COVID-19 pandemic. Decreased disease severity by the wearer has also been linked to the use of cloth facemasks. This well-documented finding is surprising considering that such masks are poor at filtering the smallest aerosol particles, which can reach the lower respiratory tract and have been associated with severe disease. We show that facemasks strongly increase the effective humidity of inhaled air, thereby promoting hydration of the respiratory epithelium which is known to be beneficial to the immune system. Increased humidity of inspired air could be an alternate explanation for the now well-established link between mask wearing and lower disease severity.

## INTRODUCTION

Respiratory viral infections are perhaps the most common type of illness. They range from influenza, colds, and measles to, most recently, COVID-19. The common cold encompasses more than 200 different viruses [1], including a large family of rhinoviruses, but also members of the coronavirus family, including 229E, NL63, OC43, and HKU1, that are usually associated with mild to moderate upper-respiratory tract illness. As suggested by their “common cold” name, these diseases have a seasonal character, and most of these viruses become more widespread with colder outside temperature. Person-to-person transmission of respiratory viruses is dominated by respiratory droplets generated by the infected person, which include breath, speech, cough, and sneeze droplets [2]. The types of droplets that dominate the transmission path depend on the virus and on the location of the respiratory tract infection[2].

The seasonality of COVID-19 [3] is now increasingly accepted as an important factor in the rapid escalation of this disease in the northern hemisphere with the onset of the 2020 fall and the approaching winter [4]. This pattern follows the same trend reported for the OC43 and 229E members of the coronavirus family by Kim et al., who noted that infections “sharply increased during the low temperature winter months of October through February” [4, 5]. Many factors may contribute to this seasonality. These include the following: more time spent indoors [3-5], where respiratory virus-containing aerosols remain airborne for many minutes [6-8]; decreased exposure to sunlight, resulting in lower levels of vitamin D, which is essential to the immune system [9]; lower UV levels that efficiently inactivate larger coronaviruses such as SARS-CoV-2, the causative agent of COVID-19; and prolonged viability of the virus at lower temperature and lower humidity [10-12]. Importantly, outdoor temperature is positively correlated with indoor relative humidity, which can reach low levels during the colder winter months. Because lower relative humidity results in faster dehydration of respiratory droplets, a larger fraction of droplets fully dehydrate before landing on the ground [13]. Hence, the fraction that remains as aerosol, and thereby the potential for transmission, increases. All of these factors likely play some role in the seasonality of respiratory viruses and can be considered “external factors”, impacting the life cycle of the virus outside of the human host.

Other factors that relate seasonality to disease concern how the host reacts to the viral infection. Iwasaki and co-workers demonstrated in mice that low humidity increases disease severity following a respiratory challenge with the influenza A virus [14]. This effect was attenuated in caspase-1/11–deficient Mx1 mice and was linked to diminished interferon-stimulated gene expression in response to the viral infection, thereby impairing the innate antiviral defense. Dehydration of the airways upon inspiration of low absolute humidity air results in respiratory water loss that makes the surface layer hyperosmolar. This elevated osmolarity causes extraction of water from the underlying epithelial cells, which decreases their volume and causes the airways to shrink, an effect exacerbated in exercise-induced asthma [15]. Dehydration of the airways is also known to result in decreased mucociliary clearance of pathogens from the lungs [16, 17].

Small droplets generated by breathing (0.3-2 μm diameter) have also been proposed to serve as a vehicle in spreading the virus through the lower respiratory tract by self-inoculation [18]. Despite their small size, such droplets are still one to three orders of magnitude larger in volume than the SARS-CoV-2 virus and therefore can easily encapsulate one or more virions. Moreover, recent work indicates a strong increase in droplet count upon SARS-CoV-2 infection of the lungs of non-human primates [19], as well as high levels of viral shedding in the exhaled breath of hospitalized patients [20].

Breath droplet formation results from transient occlusion of the small airways that can occur upon expiration [21, 22]. Subsequent inspiration then results in a thin film in the occluded airway just before it bursts open, a droplet-generating process that depends on the surface tension, viscosity, and hydration of the film fluid. This fluid, which can contain virus, derives from the ca 1-micron thick mucosal air-surface layer that floats on top of the less viscous cilia-containing serous layer. Hence, breath droplet generation, and thereby virus aerosolization, is impacted by the hydration state of the epithelial surface [21, 22].

Bromhexine, an over-the-counter medication, has shown remarkably positive results for limiting disease severity in recent clinical trials [23, 24]. Although the drug was postulated to function as an inhibitor of the human TMPRSS2 protease [24], thereby blocking requisite cleavage of the SARS-CoV-2 Spike protein, its affinity for TMPRSS2 (K_d_ ∼0.7 μM) appears too low for this inhibition to be effective. Instead, we propose its benefits may derive from its ability to enhance mucociliary clearance through hydrolytic depolymerization of mucus protein fibers as well as increased hydration of the epithelial surface, which both relate to its use in general clinical practice as a secretolytic expectorant. Interestingly, the anti-asthma drug budesonide, used as an inhaled rather than a systemic corticosteroid [25], also is showing remarkably positive preliminary results in its clinical trial (STOIC [26]). Although its anti-COVID-19 mechanism remains under investigation, its natural mode of action relates to keeping the small airways open, thereby reducing breath droplet generation.

Although much debate remains about the relative importance of the various season-related factors on the transmissibility and severity of the COVID-19 disease, the correlation between increased disease severity and low humidity of inhaled air appears strong. Equally striking are recent reports that link the use of face coverings to reduced disease severity in wearers of generic face masks [27, 28], but the authors’ suggested mechanism that reduced dose of virus is responsible for the lower severity remains contested [29].

We propose that the attenuating effect of face masks on coronavirus-2019 (COVID-19) disease severity [27, 28] is dominated by the substantial increase in the effective humidity of inspired air, where the mask acts as a temporary water storage site. The mask absorbs much of the water in exhaled breath that becomes super-saturated upon cooling when exiting the mouth; upon subsequent inspiration of dry air, this water evaporates and thereby humidifies the air that passes through this hydrated mask. We measured the magnitude of the effect at temperatures ranging from 8 to 37 °C, and for different types of mask material. Whereas all masks tested result in substantial humidification, the effect is strongest for high density cotton masks, where the high heat capacity of such masks aids to heat and humidify the inspired air, resulting in effective increases above the environmental humidity that can exceed 100%. Hence, such masks act as rudimentary equivalents to the more effective heat-exchanger masks, introduced decades ago to mitigate cold-induced asthma [30, 31].

## METHODS

Measurements were made by breathing into a sealed steel box with dimensions 38.74 × 38.74 × 63.50 cm (width/height/depth), for a total volume of 95.3 L (SI Figure S1). The side walls of the chamber were factory-powder-coated, resulting in minimal moisture adherence. The rear wall, originally bare steel, was oil-base painted for minimal moisture adherence. The high thermal conductivity and heat capacity of the box reduced temperature increases upon expiration of warm breath into the chamber to less than 0.6 °C. The acrylonitrile-butadiene-styrene front panel contains a sealable opening, shaped to accommodate the chin, mouth, and nose of the breathing volunteer, with the edges of the opening lined with high density foam rubber to make a tight seal with the facial skin of the volunteer. The front panel also contains a second, sealable, 10 cm diameter hole for refreshing the contents of the box with environmental air by using a high capacity (300 L/min) fan mounted in front of this opening between measurements, with the breathing opening serving as the channel for air exhaust. Sensors to record temperature, humidity, and CO_2_ levels were mounted close to the geometric center of the box. A 10-cm fan (flow rate 100 L/min), positioned at the bottom of the box and pointed upward at a 45° angle, served to keep the contents of the box homogeneous and to maximize airflow over the three types of sensors. The response to a step change in humidity was found to depend on temperature (SI Figure S2), defined by exponential time constants of 15, 5, and 2.8 s at 8, 22, and 37 °C, respectively. At the top rear end of the box, an inflatable polyethylene bag was mounted to accommodate the changes in total gas volume at ambient pressure upon breathing. This bag also served as a visual gage to control the volume of the exhaled air.

All data reported here were recorded under conditions aimed to simulate tidal breathing at a rate of 10 breaths per minute, and a volume of 0.95 L per breath. Although this volume is 10-50% higher than often used for tidal breathing, it was adjusted for optimal comfort of the volunteer and accounts for the decreased O_2_ uptake when the CO_2_ level in the chamber increased to >6000 ppm during the breathing maneuver. The breathing was synchronized with an audible timer, and relative humidity and temperature data were monitored by video and manually entered into a computer for analysis. Under each condition, each measurement was repeated three times, with raw values reported in SI Table S1.

Measurements were carried out in a laboratory cold room, at 7.7 ± 0.1 °C, with dry air (relative humidity (RH) 13 ± 0.5%) used to vent the box prior to measurements; indoors at room temperature (22.1 ± 0.1 °C; RH 26.6 ± 0.3%); and in a cell culture room at 37.4 ± 0.5 °C; RH 14.3 ± 0.3%. Four types of masks materials were evaluated: N95 (3M, model 9205); standard, disposable 3-ply surgical (NIH stockroom); a two-ply cotton-polyester blend mask (NIH stockroom); and an online-purchased all-cotton mask (SI Figure S3). Results were compared to the humidification of the box interior obtained for the same breathing maneuver without a mask.

Relative humidity was measured using a Honeywell HIH-5030 humidity sensor powered by a regulated 5-V power supply. The measured voltages were converted to relative humidity using the formulas given in the sensor’s datasheet. Relative humidity (RH, %) measurements were converted to absolute humidity (AH, mg/L) values according to:

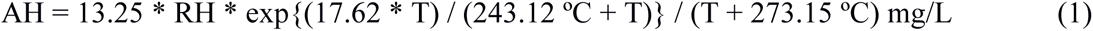

where T is the temperature in °C. Eq. 1 was derived from the ideal gas law using the empirical equation for predicting the saturation vapor pressure of water from the Guide to Meteorological Instruments and Methods of Observation, World Meteorological Organization, 2008 [32]. The change in mass of water vapor in the measurement chamber was calculated from the difference in absolute humidity, measured before and after breathing, multiplied by 95.3 L, the volume of the chamber. The amount of water absorbed by each mask, *m*_abs_(H_2_O)_mask_, was calculated by subtracting the mass of water vapor added to the chamber when wearing the mask, Δ*m*_chamber_(H_2_O)_mask_, from the mass added when not wearing the mask, Δ*m*_chamber_(H_2_O)_none_:

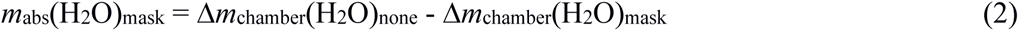

For mixing a single breath with the contents of the chamber, the absolute humidity after this breath is given by,

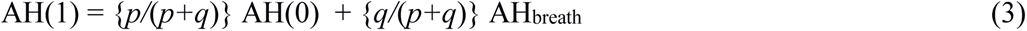

where *p* is the volume of the chamber, *q* is the volume of exhaled breath, and AH(*n*) is the absolute humidity of the chamber after *n* breaths. Recursive application of Eq. 3 to account for the effect of *n* successive breaths yields:

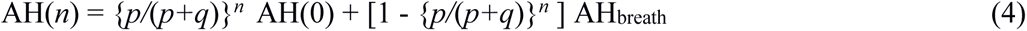

Provided *p* and *q* are known, the absolute humidity of exhaled breath is then obtained from

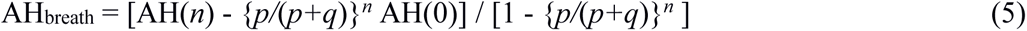

The above equations assume the air is well mixed, aided by the fan mounted inside the chamber, by the time the air is inspired from the chamber. At 22 °C, the absolute humidity of exhaled breath obtained using Eq. 5 corresponded to 38.6 ± 2 mg/L, within experimental uncertainty of the value of 37.5 mg/L expected for breath emitted at 35 °C, 95% RH, and within the range observed previously by a variety of methods [33].

The mass of water, *m*_abs_(H_2_O)_mask_, stored and released in the mask during *n* breathing cycles, corresponds to

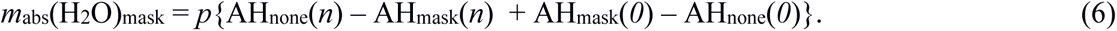

Knowing the volume of each breath, *q*, and the number of breathing cycles, the increase in absolute humidity of inspired air over the average absolute humidity in the chamber is given by *m*_abs_(H_2_O)_mask_/(*rq*), where *r* is the number of breath cycles, and *m*_abs_(H_2_O)_mask_ is converted to an increase in inspired RH by using Eq. 1 at the average temperature of the chamber during the measurement.

## RESULTS

The current measurements of the effect of masks on the humidity of inhaled air were motivated by observations made while testing various types of face coverings for blocking speech and breath particles, using laser light scattering observation [7, 34, 35]. During those measurements, the volume of exhaled air was simply derived from the increase in relative humidity within the chamber, which proved to be a convenient method in the absence of masks. However, it became obvious that considerably more expiration/inspiration breathing cycles were needed to achieve the same increase in RH with a mask than without a mask. This observation indicated that the mask absorbs water from the exhaled breath, which under steady-state conditions must be released upon inspiration, thereby effectively increasing the humidity of inhaled air. Considering the increasingly recognized impact of humidity on the spread and severity of respiratory diseases, including COVID-19, we quantitatively measured the effective increase in the humidity of inspired air when using facemasks.

Measurements were carried out at three different temperatures, from 8 to 37 °C, which covers the most relevant range from a work and living environment perspective. The ability of various types of synthetic and natural fibers to absorb water depends strongly on temperature [36-38]. Natural fibers such as wool, cotton, and silk are particularly effective at absorbing water, whereas synthetic polyester or nylon fibers do so to a much lesser extent [38]. In our pilot study, we simply tested four common types of masks: An N95 respirator mask; a regular surgical mask; an NIH-supplied mask consisting of two-layers of a blend of cotton and polyester; and a relatively thick, lined cotton mask with a total mass of 22 g (Figure S3). In all cases, leakage around the edges of the mask was eliminated by the tight fit of the volunteer’s face against the high-density foam rubber surrounding the opening in the front panel used for breathing (Figure S1).

For testing, the temperature and humidity of the mask were equilibrated on the face of the volunteer for at least 10 minutes prior to the start of the measurements. After a few cycles of optimizing and controlling the rate and quantity of breathing, measurements proved highly reproducible and self-consistent. In particular for the measurements at room temperature, where it was possible to keep room temperature and humidity within a very narrow range, the three repeats of each measurements were frequently within the digital readout of the humidity sensor, corresponding to 0.3% change in RH (Figure 1).

**FIGURE 1.**
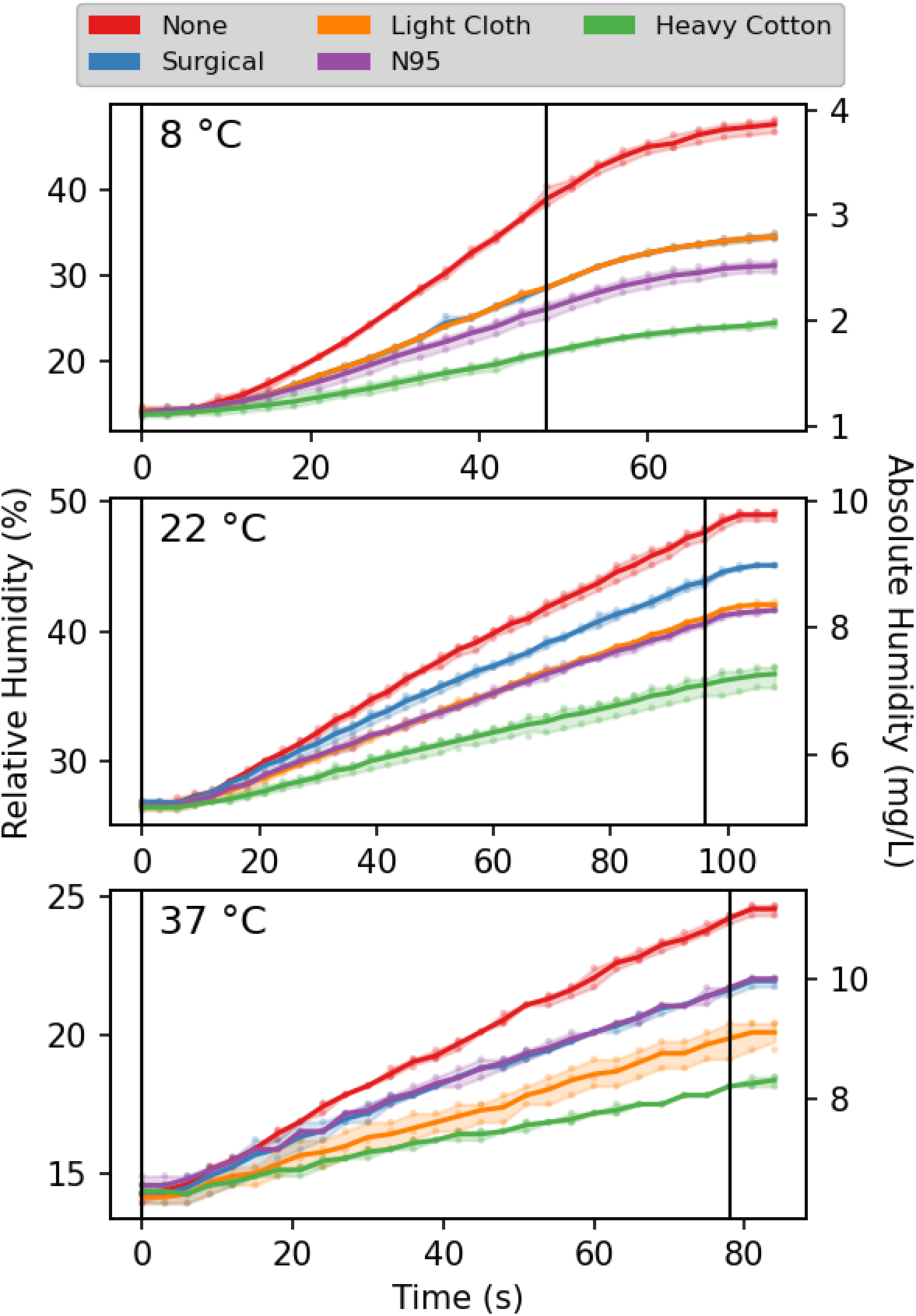
Humidity in the breathing chamber, during tidal breathing at a rate of ten 0.95-L breaths per minute without and with facemasks. The absolute humidity is derived from the relative humidity sensor readings, assuming that the total temperature increases less than 0.5 °C for all measurements, recorded at 0.1 °C resolution, is linear in time. Solid lines connect the mean of three measurements, taken at each time point. The shaded areas enclose the 95% confidence intervals. Black vertical lines mark the start and end of breathing. The temperature-dependent delay in response time of the humidity sensor is evident in the first few seconds of the trajectories and the final asymptotic stabilization after the end of breathing. The lag is most pronounced at low temperature (SI Figure S2).

All four different types of mask were found to strongly decrease the build-up of humidity in the chamber with breathing, but to different extents. At room temperature, the smallest increase in apparent RH of inspired air over the average RH of the chamber by *ca* 35% was observed for the surgical mask. Under the same conditions, both the N95 mask and the polyester-cotton mask increased the humidity of inspired air by 55-60% (Figure 2). Surprisingly, measurements with the heavy cotton mask resulted in a strong further decrease of chamber humidification, meaning a considerably larger fraction of exhaled water is temporarily stored in the fabric mask. During the 96-second measurement, the apparent RH increase over the average RH (*ca* 30%) in the chamber was about 90%. At first sight, this apparent increase of inspired air humidity to above 100% may defy intuition. However, the explanation is grounded in simple physics. The exhalation of breath warms up the mask (and the water stored in it) to well above the environmental temperature. Indeed, with the air temperature at 8 °C, rapidly removing the mask after exhalation during tidal breathing, folding it double while clamping a thermocouple sensor near its center-fold, followed by rolling up the doubled mask, consistently yielded temperature readings of 30±1 °C. The same measurement after inspiration yielded 27±1 °C. Presumably, this 30±1 °C temperature represents a lower limit for the actual temperature at the face side of the mask at the start of inspiration, allowing the humidity of inspired air to reach values that are well above 100% at 8 °C. Even if only 20% of the mask surface was involved in the actual passage of air, this fraction of the mask corresponds to about 4 grams of cotton with a total heat capacity of 20 J/K, considerably higher than the heat capacity of *ca* 1.2 J/K for one liter of inspired air. Hence, the high heat capacity of the mask causes it to function like a rudimentary heat and humidity exchanger, warming and humidifying the inspired air to values much closer to those in the lower respiratory tract than of room air.

**FIGURE 2.**
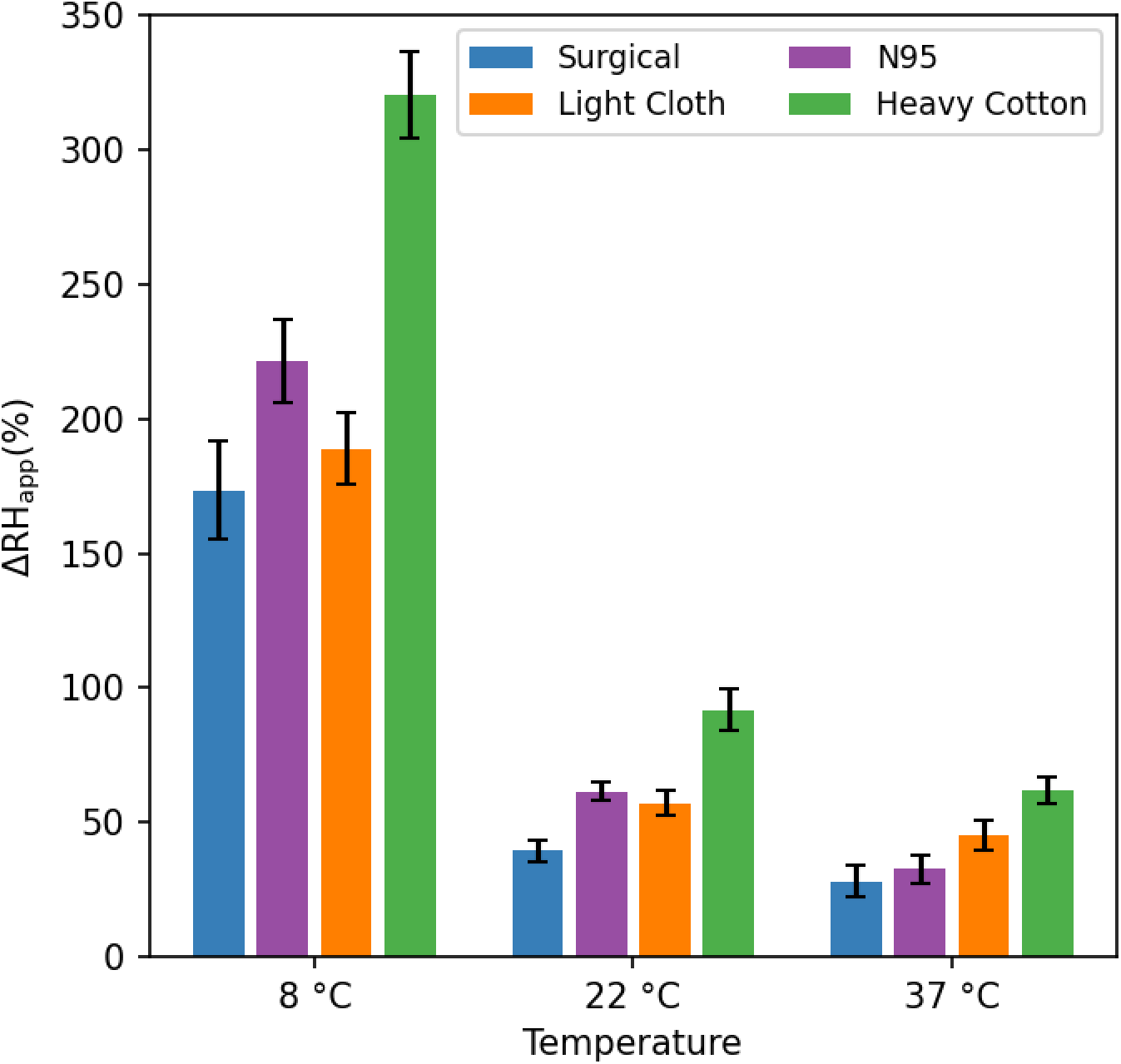
Increase in apparent relative humidity of inspired air, ΔRH_app_, during pseudo-tidal breathing for four different masks. ΔRH_app_ is derived from the increase in absolute humidity of air inspired through the mask, assuming the temperature of inhaled air is that of the room in which the measurements were carried out, and does not account for the increase, ΔT, in temperature of the gas when traversing the mask, which is particularly large for the heavy cotton mask (ΔT ≈ 22±2 °C at 8 °C; 8±1 °C at 22 °C; 0 °C at 37 °C) and enables >100% RH increases over that of the room air.

The considerably lower increases seen for the lighter masks are consistent with this general concept, although the filtering material itself also appears to play a significant role. For example, despite its low mass of *ca* 27 mg/cm^2^, the N95 mask increases the humidity of inspired air comparably to the ca two-fold heavier, polyester-cotton mask. It seems likely that the large surface area of the extremely fine fibers in N95 masks that carry out most of the actual filtering [39], together with their electrostatic properties, is responsible for their ability to absorb a substantial fraction of the exhaled water vapor.

At low temperature (8 °C), the humidifying effect of all masks strongly increases. It is important to note, however, that these very large RH increases at 8 °C (150-300%) correspond to less dramatic increases in absolute humidity of 12-24 mg/L. Once heated inside the respiratory tract, this corresponds to changes in relative humidity at 36 °C of 29-58%. By contrast, even while the RH changes of inspired air are much smaller for the measurements at 37 °C, this increase directly translates into the relative humidity change of air inspired into the lungs. As a consequence, from the perspective of respiratory tract dehydration, the effect of each mask type is fairly constant across the entire 8-37 °C range. At all temperatures, the humidification resulting from the heavy cotton mask is about double that of a surgical mask, with the N95 and cotton-polyester cloth masks falling in between.

## DISCUSSION

Over the past year, it has become abundantly clear that facemasks offer an effective tool for containing the spread of the COVID-19 pandemic. Remarkably, mask usage correlated not only a protective effect in terms of disease transmission but also to a striking decrease in disease severity, the latter effect attributed to the infected person receiving a lower dose of the disease-causing virus [27]. The effect of masks was so pronounced that the authors even raised the possibility of “variolation”, i.e., the inoculation of an uninfected person with a minute amount of live virus, a procedure introduced into the Western world about three centuries ago to fight smallpox (*Variola*) prior to the availability of a vaccine [28]. Although, following the successful development of multiple effective COVID-19 vaccines, the idea of variolation is no longer relevant and was strongly criticized by experts [29], we also note that it should not apply to airborne viral respiratory diseases in the first place. As shown by Haas et al. [40], the dose-response curve for SARS infection follows the exponential model, also known as Single-Hit-Model (SHM) [41] or Independent Action Hypothesis (IAH) [42], meaning that the risk of infection initially increases linearly with exposure. This model also proved appropriate in the classic analysis of a measles outbreak [8]. Even while any single inhaled virion is unlikely to enter a susceptible host cell and create progeny, the probability of such an infection to occur is non-zero and increases linearly with the number of virions inhaled. This is analogous to purchasing lottery tickets: the odds of a winning single ticket are small but increase linearly with the number of tickets. With virions randomly dispersed in the oral or mucosal fluid that exits the mouth of an infected person in the form of highly hydrated (>95%) microscopic droplets, the vast majority of these droplets are so small that statistically they are unlikely to contain more than one infectious virion [7]. In contrast to smallpox *variolation*, where the virus was applied to a small scratch on the skin, inhaled airborne SARS-CoV-2 virions will be distributed randomly across the epithelial surface of the respiratory tract. This makes any requirement for overload of the innate immune system, a concept that applies locally, highly unlikely.

There is a second problem with the proposal that reduced COVID-19 disease severity of mask wearers results from lower exposure. It is well recognized that only particles smaller than a few microns can enter the small airways and cause infection of the lungs, commonly associated with *increased* disease severity relative to infection of the upper respiratory tract [2, 43, 44]. However, cloth masks are relatively poor at filtering out these smallest particles. So, even though cloth masks are expected to lower the incidence of infection, their protective effect to the wearer will be less for the smallest particles, which are associated with more severe disease, opposite to observation. An alternate explanation for the attenuating effect of masks on disease severity is therefore needed.

Here, we propose that the increased humidity of air inspired through face masks is responsible for the lower disease severity of mask wearers [27, 28]. In contrast to the primary function of masks to prevent virus from entering the respiratory tract of another person [45, 46], this additional benefit applies after a virus-containing particle lands on the surface of the respiratory tract: reduced dehydration limits impairment of the innate immune system [14] while improving mucociliary clearance [16, 17], with both these factors reducing infection probability. If an infection does occur, humidification may limit its further spread through the lungs by lowering the generation of virus-containing breath droplets that could lead to self-inoculation elsewhere in the lungs [18, 19]. At the same time, effective mucociliary clearance and an unimpaired innate immune system of the well-humidified respiratory tract also may limit viral spreading, allowing more time for mobilization of the adaptive immune system.

The increased humidity of inspired air associated with wearing a face cover is perhaps well recognized by the public and contributes to the general feeling of mugginess, in particular when the weather is humid. Our measurements confirm that the increased humidity of inspired air is real and quite large.

It is important to note that our measurements were carried out in the absence of air leakage around the mask edges. For non-N95 masks, such leakage is often significant and will proportionately lower the effective humidity increase of inspired air. So, in practice the cotton mask humidification efficiency will drop somewhat, to values comparable to the tight fitting N95 mask. The surgical and polyester-cotton blend masks, which also will be subject to leakage, then are expected to perform somewhat lower than the N95 and all-cotton masks.

## Data Availability

All data acquired by the authors and details of the experimental setup are available from the authors upon request

## AUTHOR INFORMATION

### Notes

The authors declare no competing financial interests.

### Author Contributions

A.B. and J.M.C. designed research; A.B. and J.M.C. performed research; J.M.C. analyzed data A.B. and J.M.C. wrote the paper.

## Acknowledgments

We thank Philip Anfinrud, Ingrid Pufahl, Dennis Torchia, William Eaton, Kevin Fennelly, Dan Nicolau and Mona Bafadhel for valuable discussions. This work was supported by the Intramural Research Program of the National Institute of Diabetes and Digestive and Kidney Diseases and the NIH Intramural Antiviral Target Program.

## References

1. Wein, H., Understanding a Common Cold Virus. https://www.nih.gov/news-events/nih-research-matters/understanding-common-cold-virus, 2009.

2. Musher, D.M., Medical progress: How contagious are common respiratory tract infections? N. Engl. J. Med., 2003. 348(13): p. 1256–1266.

3. Merow, C. and M.C. Urban, Seasonality and uncertainty in global COVID-19 growth rates. Proc. Natl. Acad. Sci. U. S. A., 2020. 117(44): p. 27456–27464.

4. Audi, A., M. AlIbrahim, M. Kaddoura, G. Hijazi, H.M. Yassine, and H. Zaraket, Seasonality of Respiratory Viral Infections: Will COVID-19 Follow Suit? Frontiers in Public Health, 2020. 8.

5. Kim, J.M., J.S. Jeon, and J.K. Kim, Climate and Human coronaviruses 229E and Human coronaviruses OC43 Infections: Respiratory Viral Infections Prevalence in Hospitalized Children in Cheonan, Korea. Journal of Microbiology and Biotechnology, 2020. 30(10): p. 1495–1499.

6. Fennelly, K.P., Particle sizes of infectious aerosols: implications for infection control. Lancet Respiratory Medicine, 2020. 8(9): p. 914–924.

7. Stadnytskyi, V., C.E. Bax, A. Bax, and P. Anfinrud, The airborne lifetime of small speech droplets and their potential importance in SARS-CoV-2 transmission. Proc. Natl. Acad. Sci. USA, 2020: p. 202006874.

8. Riley, E.C., G. Murphy, and R.L. Riley, AIRBORNE SPREAD OF MEASLES IN A SUBURBAN ELEMENTARY SCHOOL. American Journal of Epidemiology, 1978. 107(5): p. 421–432.

9. Grant, W.B., H. Lahore, S.L. McDonnell, C.A. Baggerly, C.B. French, J.L. Aliano, and H.P. Bhattoa, Evidence that Vitamin D Supplementation Could Reduce Risk of Influenza and COVID-19 Infections and Deaths. Nutrients, 2020. 12(4).

10. Chin, A.W.H., J.T.S. Chu, M.R.A. Perera, K.P.Y. Hui, H.-L. Yen, M.C.W. Chan, M. Peiris, and L.L.M. Poon, Stability of SARS-CoV-2 in different environmental conditions. Lancet Microbe, 2020. 1(1): p. e10–e10.

11. Dabisch, P., M. Schuit, A. Herzog, K. Beck, S. Wood, M. Krause, D. Miller, W. Weaver, D. Freeburger, I. Hooper, B. Green, G. Williams, B. Holland, J. Bohannon, V. Wahl, J. Yolitz, M. Hevey, and S. Ratnesar-Shumate, The influence of temperature, humidity, and simulated sunlight on the infectivity of SARS-CoV-2 in aerosols. Aerosol Sci. Technol.

12. Matson, M.J., C.K. Yinda, S.N. Seifert, T. Bushmaker, R.J. Fischer, N. van Doremalen, J.O. Lloyd-Smith, and V.J. Munster, Effect of Environmental Conditions on SARS-CoV-2 Stability in Human Nasal Mucus and Sputum. Emerging Infectious Diseases, 2020. 26(9): p. 2276–2278.

13. Netz, R.R. and W.A. Eaton, Physics of virus transmission by speaking droplets. Proceedings of the National Academy of Sciences, 2020. 117(41): p. 25209–25211.

14. Kudo, E., E. Song, L.J. Yockey, T. Rakib, P.W. Wong, R.J. Homer, and A. Iwasaki, Low ambient humidity impairs barrier function and innate resistance against influenza infection. Proc. Natl. Acad. Sci. U. S. A., 2019. 116(22): p. 10905–10910.

15. Anderson, S.D. and E. Daviskas, The mechanism of exercise-induced asthma is. Journal of Allergy and Clinical Immunology, 2000. 106(3): p. 453–459.

16. Williams, R., N. Rankin, T. Smith, D. Galler, and P. Seakins, Relationship between the humidity and temperature of inspired gas and the function of the airway mucosa. Critical Care Medicine, 1996. 24(11): p. 1920–1929.

17. Wolkoff, P., The mystery of dry indoor air - An overview. Environment International, 2018. 121: p. 1058–1065.

18. Edwards, D.A., J.C. Man, P. Brand, J.P. Katstra, K. Sommerer, H.A. Stone, E. Nardell, and G. Scheuch, Inhaling to mitigate exhaled bioaerosols. Proc. Natl. Acad. Sci. U. S. A., 2004. 101(50): p. 17383–17388.

19. Edwards, D.A., D. Ausiello, R. Langer, J. Salzman, T. Devlin, B.J. Beddingfield, A.C. Fears, L.A. Foyle-Meyers, R.K. Redmann, S.Z. Killeen, N.J. Maness, and C.J. Roy, Exhaled aerosol increases with COVID-19 infection,and risk factors of disease symptom severity. https://doi.org/10.1101/2020.09.30.20199828 doi: medRxiv preprint, 2020.

20. Ma, J., X. Qi, H. Chen, X. Li, Z. Zhang, H. Wang, L. Sun, L. Zhang, J. Guo, L. Morawska, S.A. Grinshpun, P. Biswas, R.C. Flagan, and M. Yao, COVID-19 patients in earlier stages exhaled millions of SARS-CoV-2 per hour. Clin Infect Dis, 2020: p. ciaa1283.

21. Johnson, G.R. and L. Morawska, The Mechanism of Breath Aerosol Formation. Journal of Aerosol Medicine and Pulmonary Drug Delivery, 2009. 22(3): p. 229–237.

22. Bake, B., P. Larsson, G. Ljungkvist, E. Ljungstrom, and A.C. Olin, Exhaled particles and small airways. Respiratory Research, 2019. 20.

23. Depfenhart, M., D. de Villiers, G. Lemperle, M. Meyer, and S. Di Somma, Potential new treatment strategies for COVID-19: is there a role for bromhexine as add-on therapy? Intern Emerg Med, 2020. 15(5): p. 801–812.

24. Ansarin, K., R. Tolouian, M. Ardalan, A. Taghizadieh, M. Varshochi, S. Teimouri, T. Vaezi, H. Valizadeh, P. Saleh, S. Safiri, and K.R. Chapman, Effect of bromhexine on clinical outcomes and mortality in COVID-19 patients: A randomized clinical trial. Bioimpacts, 2020. 10(4): p. 209–215.

25. Nicolau, D.V. and M. Bafadhel, Inhaled corticosteroids in virus pandemics: a treatment for COVID-19? Lancet Respiratory Medicine, 2020. 8(9): p. 846–847.

26. Bafadhel, M. and D.V. Nicolau, Evaluate the effect of intervention on emergency department attendance or hospitalisation related to COVID-19, https://www.clinicaltrials.gov/ct2/show/record/NCT04416399, Editor. 2020.

27. Gandhi, M., C. Beyrer, and E. Goosby, Masks Do More Than Protect Others During COVID-19: Reducing the Inoculum of SARS-CoV-2 to Protect the Wearer. Journal of General Internal Medicine, 2020.

28. Gandhi, M. and G.W. Rutherford, Facial Masking for Covid-19 — Potential for “Variolation” as We Await a Vaccine. N. Engl. J. Med., 2020. 383(18): p. e101.

29. Brosseau, L.M., C.J. Roy, and M.T. Osterholm, Facial Masking for Covid-19. N. Engl. J. Med., 2020. 383(21): p. 2092–2094.

30. Nisar, M., D.P.S. Spence, D. West, J. Haycock, Y. Jones, M.J. Walshaw, J.E. Earis, P.M.A. Calverley, and M.G. Pearson, A mask to modify inspired air-temperature and humidity and its effect on exercise induced asthma. Thorax, 1992. 47(6): p. 446–450.

31. Beuther, D.A. and R.J. Martin, Efficacy of a heat exchanger mask in cold exercise-induced asthma. Chest, 2006. 129(5): p. 1188–1193.

32. Organization, W.M. Guide to Meteorological Instrumentsand Methods of Observation. http://www.posmet.ufv.br/wp-content/uploads/2016/09/MET-474-WMO-Guide.pdf page I.4-29. 2020 [cited 2020 December 13].

33. Morawska, L., G.R. Johnson, Z.D. Ristovski, M. Hargreaves, K. Mengersen, S. Corbett, C.Y.H. Chao, Y. Li, and D. Katoshevski, Size distribution and sites of origin of droplets expelled from the human respiratory tract during expiratory activities. J. Aerosol Sci., 2009. 40(3): p. 256–269.

34. Anfinrud, P., V. Stadnytskyi, C.E. Bax, and A. Bax, Visualizing Speech-Generated Oral Fluid Droplets with Laser Light Scattering. N. Engl. J. Med., 2020.

35. Fischer, E.P., M.C. Fischer, D. Grass, I. Henrion, W.S. Warren, and E. Westman, Low-cost measurement of face mask efficacy for filtering expelled droplets during speech. Science Advances, 2020. 6(36).

36. Ashpole, D.K., The moisture relations of textile fibres at high humidities. Proc. Roy. Soc. Math. Phys. Sci., 1952. 212(1108): p. 112–123.

37. Toner, R.K., C.F. Bowen, and J.C. Whitwell, Equilibrium moisture relations for textile fibers. Textile Res. J., 1947. 17(1): p. 7–18.

38. International, A., ASTM D1909 - 13(2020)e1 Standard Tables of Commercial Moisture Regains and Commercial Allowances for Textile Fibers. 2018, https://www.astm.org/Standards/D1909.htm

39. Lee, H.R., L. Liao, W. Xiao, A. Vailionis, A.J. Ricco, R. White, Y. Nishi, W. Chiu, S. Chu, and Y. Cui, Three-Dimensional Analysis of Particle Distribution on Filter Layers inside N95 Respirators by Deep Learning. Nano Lett., 2020.

40. Watanabe, T., T.A. Bartrand, M.H. Weir, T. Omura, and C.N. Haas, Development of a Dose-Response Model for SARS Coronavirus. Risk Analysis, 2010. 30(7): p. 1129–1138.

41. Meynell, G.G. and B.A.D. Stocker, Some hypotheses on the aetiology of fatal infections in partially resistant hosts and their application to mice challenged with salmonella-paratyphi-B or salmonella-typhimurium by intraperitoneal injection. J. Gen. Microbiol., 1957. 16(1): p. 38–58.

42. Zwart, M.P., J.A. Daros, and S.F. Elena, One Is Enough: In Vivo Effective Population Size Is Dose-Dependent for a Plant RNA Virus. PLoS Pathog., 2011. 7(7): p. 12.

43. Raymond, T., Review of Aerosol Transmission of Influenza A Virus. Emerging Infectious Disease journal, 2006. 12(11): p. 1657.

44. Gralton, J., E. Tovey, M.L. McLaws, and W.D. Rawlinson, The role of particle size in aerosolised pathogen transmission: A review. J. Infection, 2011. 62(1): p. 1–13.

45. Howard, J., A.L. Huang, Z., Z. Tufekci, V. Zdimal, H. van der Westhuizen, A. von Delft Price, L. Fridman, L. Tang, V. Tang, G.L. Watson, C.E. Bax, R. Shaikh, F. Questier, D. Hernandez, L.F. Chu, C.M. Ramirez, and A.W. Rimoin, Face Masks Against COVID-19: An Evidence Review.. MEDXRIV Preprints 2020, 2020040203 doi: 10.20944/preprints202004.0203.v3, 2021.

46. Edelstein, P. and L. Ramakrishnan, Report on face masks for the general public https://rs-delve.github.io/addenda/2020/07/07/masks-update.html. 2020, Royal Society Delve Report.

