## Supplemental Table and Figures for "Hydrating the Respiratory Tract: An Alternative Explanation Why Masks Lower Severity of COVID-19 Disease" (from another version of this preprint: version 1, 2020-12-24)

Joseph M. Courtney and Ad Bax

Laboratory of Chemical Physics, NIDDK,

National Institutes of Health, Bethesda, MD, 20892-0520, USA

### Supporting Material

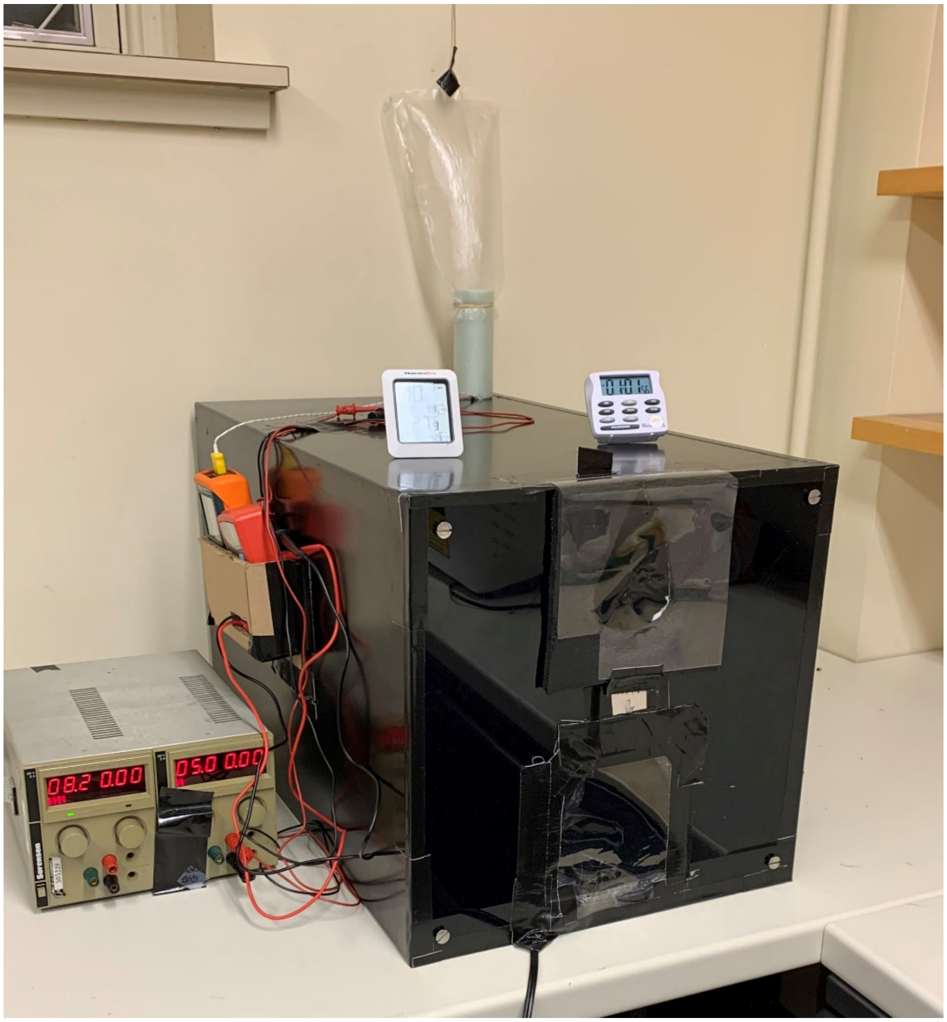

**FIGURE S1**. Breathing chamber used for humidity measurements. The steel chamber has a plastic front panel with two sealable openings. The upper opening is shaped to optimally fit the face of the volunteer and is lined with high density foam rubber to make a tight seal with the face. For purging the chamber, a fan is mounted outside the chamber in front of the bottom hole. The polyethylene bag at the top of the box is connected to the interior by a short piece of high-density pipe insulation and accommodates the change in gas volume associated with breathing into the box. Thermocouple and temperature sensor leads enter the chamber along the flexible pipe insulation.

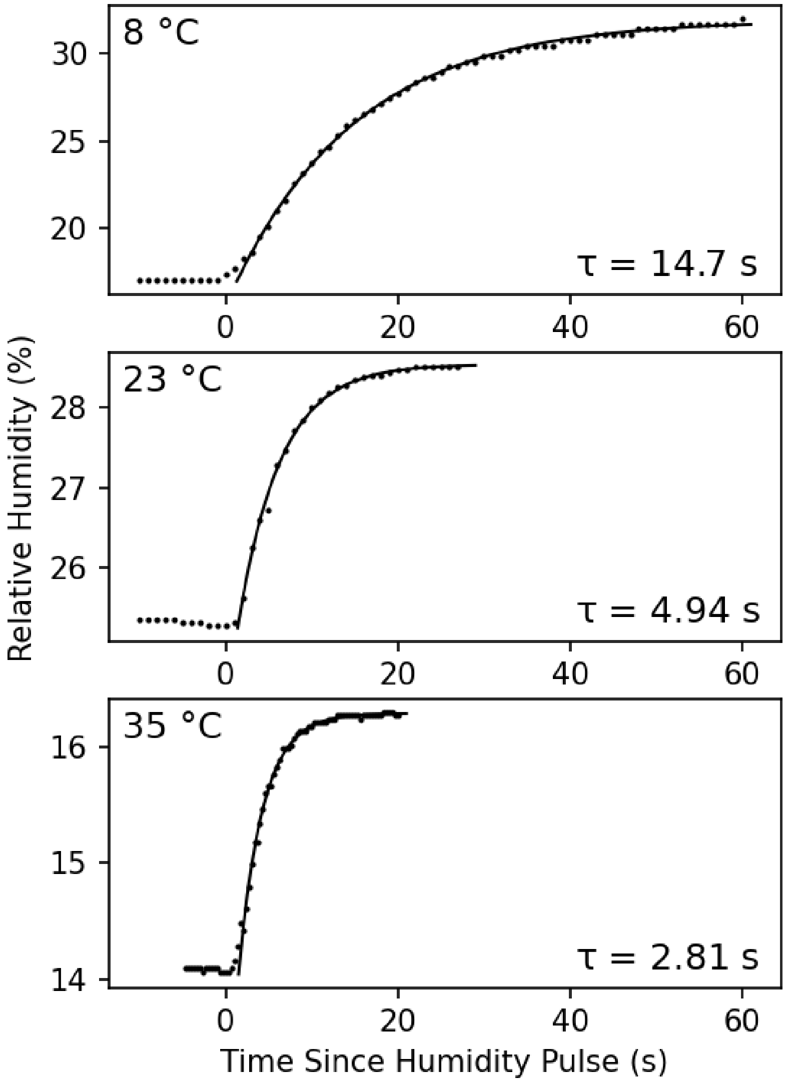


**FIGURE S2**. Humidity sensor response to a step-increase in humidity at time zero, recorded at three different temperatures. The solid lines correspond to mono-exponential fits to the step-elevated humidity of the chamber, preceded by a 1.4-s delay that is attributed to the time required for mixing of the air by the internal fan. The time constants are indicated in each panel.

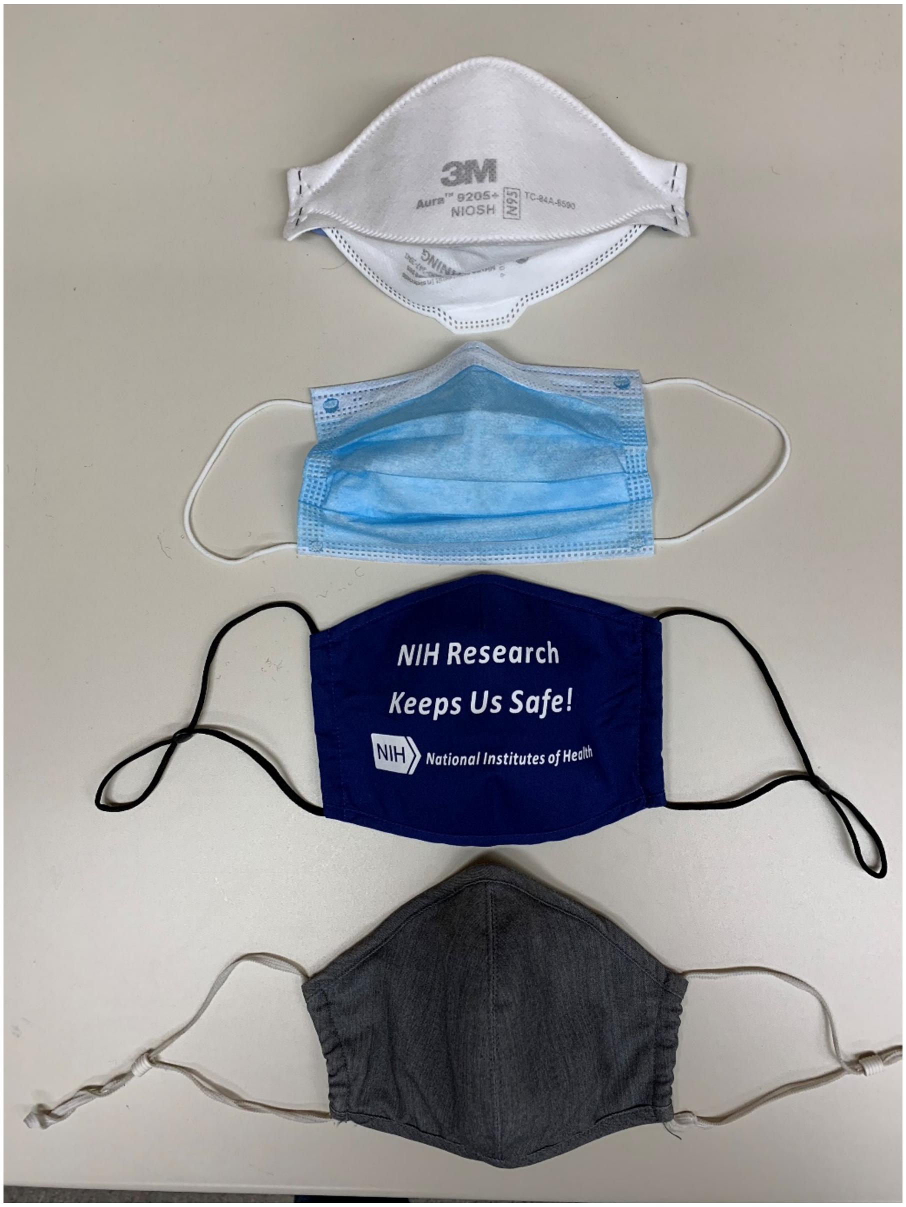


**FIGURE S3**. Four masks used for humidity measurements. From to top bottom: 3M 9205 N95 mask; NIH stockroom surgical mask; NIH stockroom polyester-cotton blend two-ply cloth mask; double-ply 100% cotton mask, purchased online from https://mandalascrubs.com/products/face-mask

**TABLE S2**. Temperature and humidity sensor readings before and after pseudo-tidal breathing. Voltages and temperatures after breathing were taken after stabilization.

| MASK | LOCATION | RUN | TEMPERATURE (°C) BEFORE | SENSOR VOLTAGE (V) BEFORE | TEMPERATURE (°C) AFTER | SENSOR VOLTAGE (V) AFTER |
|---|---|---|---|---|---|---|
| **NONE** | cold room | 1 | 7.7 | 1.20 | 7.8 | 2.35 |
| **NONE** | cold room | 2 | 7.7 | 1.20 | 7.8 | 2.30 |
| **NONE** | cold room | 3 | 7.7 | 1.20 | 7.8 | 2.35 |
| **SURGICAL** | cold room | 1 | 7.7 | 1.20 | 7.8 | 1.96 |
| **SURGICAL** | cold room | 4 | 7.7 | 1.20 | 7.6 | 1.92 |
| **SURGICAL** | cold room | 5 | 7.7 | 1.20 | 7.6 | 1.94 |
| **LIGHT CLOTH** | cold room | 1 | 7.7 | 1.20 | 7.6 | 1.91 |
| **LIGHT CLOTH** | cold room | 2 | 7.7 | 1.20 | 7.6 | 1.90 |
| **LIGHT CLOTH** | cold room | 3 | 7.7 | 1.20 | 7.8 | 1.89 |
| **N95** | cold room | 1 | 7.7 | 1.20 | 7.8 | 1.8 |
| **N95** | cold room | 2 | 7.7 | 1.20 | 7.9 | 1.81 |
| **N95** | cold room | 3 | 7.7 | 1.20 | 7.8 | 1.78 |
| **HEAVY COTTON** | cold room | 4 | 7.3 | 1.20 | 7.2 | 1.55 |
| **HEAVY COTTON** | cold room | 2 | 7.7 | 1.20 | 7.8 | 1.57 |
| **HEAVY COTTON** | cold room | 3 | 7.7 | 1.20 | 7.9 | 1.55 |
| **NONE** | office | 2 | 22.3 | 1.62 | 22.9 | 2.32 |
| **NONE** | office | 3 | 22.1 | 1.61 | 22.65 | 2.335 |
| **NONE** | office | 4 | 22.1 | 1.62 | 22.6 | 2.33 |
| **SURGICAL** | office | 1 | 22.1 | 1.62 | 22.6 | 2.20 |
| **SURGICAL** | office | 2 | 22.1 | 1.62 | 22.6 | 2.21 |
| **SURGICAL** | office | 3 | 22.1 | 1.62 | 22.4 | 2.20 |
| **LIGHT CLOTH** | office | 1 | 22.1 | 1.62 | 22.4 | 2.105 |
| **LIGHT CLOTH** | office | 2 | 22.1 | 1.61 | 22.6 | 2.11 |
| **LIGHT CLOTH** | office | 3 | 22.1 | 1.61 | 22.6 | 2.11 |
| **N95** | office | 1 | 22.1 | 1.61 | 22.6 | 2.09 |
| **N95** | office | 4 | 22.2 | 1.61 | 22.4 | 2.09 |
| **N95** | office | 5 | 22.0 | 1.61 | 22.4 | 2.09 |
| **HEAVY COTTON** | office | 2 | 21.9 | 1.61 | 22.4 | 1.95 |
| **HEAVY COTTON** | office | 3 | 22.1 | 1.61 | 22.5 | 1.90 |
| **HEAVY COTTON** | office | 4 | 22.0 | 1.60 | 22.4 | 1.95 |
| **NONE** | hot room | 1 | 38.4 | 1.20 | 38.6 | 1.53 |
| **NONE** | hot room | 2 | 37.5 | 1.20 | 37.9 | 1.52 |
| **NONE** | hot room | 3 | 37.9 | 1.20 | 37.9 | 1.52 |
| **SURGICAL** | hot room | 1 | 37.7 | 1.21 | 38.2 | 1.43 |
| **SURGICAL** | hot room | 2 | 37.6 | 1.21 | 38.2 | 1.44 |
| **SURGICAL** | hot room | 4 | 37.2 | 1.20 | 37.7 | 1.45 |

| | | | | | | |
|---|---|---|---|---|---|---|
| **LIGHT CLOTH** | hot room | 2 | 37.2 | 1.20 | 37.3 | 1.39 |
| **LIGHT CLOTH** | hot room | 3 | 37.2 | 1.20 | 37.9 | 1.37 |
| **LIGHT CLOTH** | hot room | 4 | 37.2 | 1.21 | 37.6 | 1.40 |
| **N95** | hot room | 1 | 37.4 | 1.21 | 37.6 | 1.45 |
| **N95** | hot room | 2 | 37.0 | 1.21 | 37.4 | 1.44 |
| **N95** | hot room | 3 | 37.1 | 1.20 | 37.2 | 1.44 |
| **HEAVY COTTON** | hot room | 1 | 37.0 | 1.21 | 37.5 | 1.33 |
| **HEAVY COTTON** | hot room | 2 | 37.4 | 1.20 | 37.4 | 1.33 |
| **HEAVY COTTON** | hot room | 3 | 37.5 | 1.02 | 37.7 | 1.33 |
